# Pulsed Field Ablation of Small Vessel-Related Arrhythmias: A New Catheter and Methods

**DOI:** 10.1101/2024.09.25.24314404

**Authors:** Fengqi Xuan, Yunhao Li, Jie Zhang, Long Lin, Daoyang Zhang, Qi Zhang, Ping Zhang, Yujie Zhang, Wei Ma, Zulu Wang, Yaling Han, Ming Liang

**Affiliations:** Department of Cardiology, Tianjin Chest Hospital, Tianjin, China; Department of Cardiology, General Hospital of Northern Theater Command, Shenyang, China; Graduate School of China Medical University, China Medical University, Shenyang, China; Graduate School of Dalian Medical University, Dalian Medical University, Dalian, China; National Key Laboratory of Frigid Zone Cardiovascular Diseases, Shenyang, China

## Abstract

**Objectives:** Some arrhythmia targets are located in the epicardium or deep myocardium, which could be reached through the vascular approach. However, it is difficult to deliver ablation catheters to the distal vessels and their branches. In small vessels, the energy released is limited, and the risk of ablation is increased. The objective of this study was to design a linear catheter with pulsed field energy that is suitable for ablation in the distal vessels and to verify its efficacy and safety in canines.

**Methods:** A total of eight canines were randomly assigned to two observation groups: a 48-hour group (N=4) and a 30-day group (N=4). A 3 F 10-pole pulsed field ablation catheter was employed to ablate in the great cardiac vein, the middle cardiac vein, the anterior interventricular vein, and the distal small branches of the cardiac venous system. The characteristics of the ablation lesions were observed both grossly and microscopically.

**Results:** The surgical procedure was completed successfully. Pulsed field energy can travel through the fatty tissue to form a lesion in the cardiomyocytes. The mean depth of the lesion in the 30-day group (2.37±0.53 mm) was found to be reduced by 39% (P<0.001) in comparison to that observed in the 48-hour group (3.92±0.62 mm). Additionally, a transition zone of incomplete injury was discerned in the junction area of the 48-hour group. Canines exhibited no adverse effects intraoperatively nor postoperatively, and no appreciable damage was observed in the adjacent small arteries or the surrounding organs.

**Conclusion:** The pulsed field energy from small blood vessels can cause lasting, continuous lesions to the myocardium and is safe to use. The 3 F liner pulsed field ablation catheter has been proven to be efficacious and safe, with promising indications for future application.

## 1. Background

By the endocardial approach, most arrhythmias can be successfully treated. However, there are instances that the arrhythmia targets are located in the epicardium or deep myocardium, where the ablation energy is difficult to reach. In such instances, small vessels may be chosen as optimal pathways to the targets^1,2^. Currently, arrhythmias originating from perivascular areas include 1) ventricular arrhythmias from the summit region, 2) intramural outflow tract ventricular arrhythmias, and 3) atrial arrhythmias related to Marshall’s vein, among others^3–8^. These arrhythmias are from the distal coronary vessel system and are also known as or defined as “small vessel-related ventricular arrhythmias”. Currently, the principal methods for their treatment are radiofrequency ablation, cryotherapy, chemical ablation, and guidewire ablation. However, these energy sources are subject to numerous restrictions and limited efficacy. For example, the disadvantages of radiofrequency ablation include high ablation impedance, energy release inability due to insufficient perfusion, and adjacent artery injuries. At the same time, alcohol ablation can result in non-target myocardial damage. The option of ablation energy is a significant challenge for the ablation of these arrhythmias.

Pulsed field ablation (PFA) has the advantage of high energy penetration, tissue selectivity, ignorable thermal effects, and a relatively minimal impact on adjacent arteries and conduction systems, which make it an optimal energy for intravascular ablation. A hollow 3 F pulsed field mapping and ablation catheter has been designed that can be guided by a guidewire to enter distal branches that are too small to be reached by conventional ablation catheters. Thus, the catheter can be delivered to places closer to the ablation targets anatomically. The efficacy and safety for the treatment of small vessel-related arrhythmias were confirmed in canines.

## 2. Methods and Materials

### 2.1 Animals and Materials

Eight canines were randomly assigned to either a 48-hour (N=4) or a 30-day (N=4) group and subsequently underwent surgery. Before surgery, canines were treated with dual antiplatelet therapy for 24 hours and fasted without water for 12 hours. The intravenous access was established in the cephalic vein of the forearm. Intraoperatively, they were then fixed supine, and they underwent endotracheal intubation. Anesthesia was maintained by continuous pumping of propofol. Heparin was administered by the body weight, with a target ACT of ≥300s.

A hollow 3 F 10-pole mapping and ablation liner catheter (Figure 1A, 1B) was designed and applied. The catheter had an outer diameter of 3 F and a hollow cavity diameter of 1.5 F. 10 electrodes were paired two by two. Each electrode had a width of 2 mm, a distance of 2 mm between the same pairs of electrodes, and a distance of 4 mm between electrode pairs. Pulsed field energy was applied under R-wave monitoring in a bipolar configuration between the adjacent electrodes (Electrode 1 and Electrode 3) at 1000 V by a pulse field generator 36 times (Shanghai MicroPort EP MedTech Co., Ltd, SYE-PFA-1A, Figure 1C). Each PFA application comprised multiple trains of microsecond biphasic pulses between two bipolar configurations, with a total application duration of 100 microseconds.

**Figure 1.**
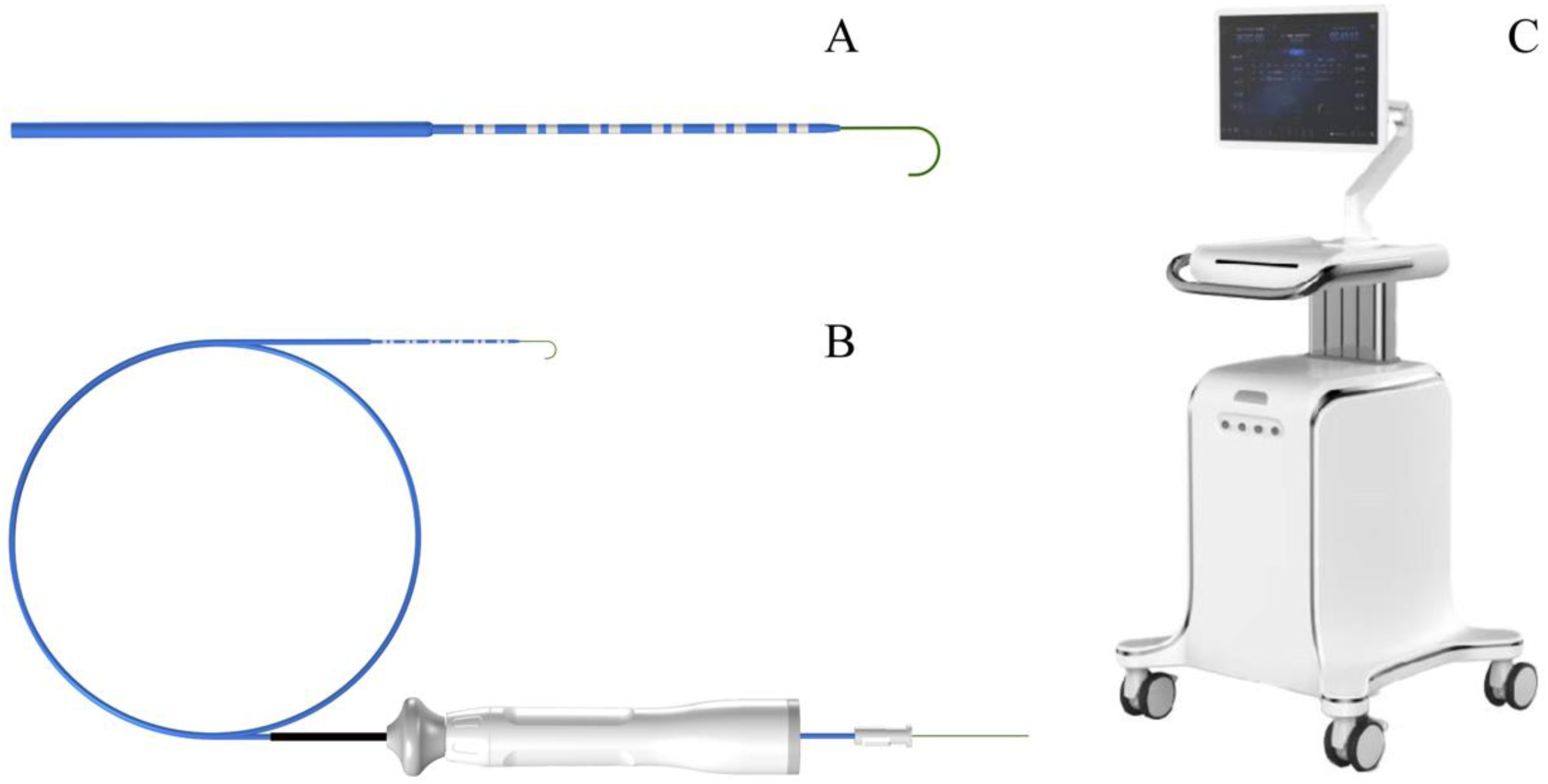
3 F guidewire-guided ten-polar pulsed field ablation mapping and ablation catheter (A, B) and the pulse field generator (C).

### 2.2 Surgical Procedure

Bilateral femoral veins were punctured, and intravenous angiography and guidewire delivery were performed via an 8.5F adjustable curved sheath (Shanghai MicroPort EP MedTech Co., Ltd). A guidewire (diameter 0.014 inches, Terumo) was introduced into the coronary venous system (CVS) under fluoroscopic via a preformed catheter (XBRCA 6F Cordis; Biosense-Webster) to reach the great cardiac vein (GCV), the middle cardiac vein (MCV), the anterior interventricular vein (AIV), and the small distal branches of the CVS. Intraoperatively, a surface electrocardiogram and an intracardiac electrocardiogram were recorded. The pericardium was monitored via DSA. Selective coronary angiography was performed before and after ablation to evaluate the pulsed field effect on adjacent arteries and to assess coronary perfusion, stenosis, or injury.

### 2.3 Postoperative Management

Canines were euthanized at 48 hours and 30 days post-surgery, and the appearance of the ablation area was observed visually. Tissue samples were excised at 5 mm intervals perpendicular to the CVS, stained with a 2% TTC solution, and incubated at 37°C for 30 minutes before gross observation. The maximum and minimum lesion depths were measured at the cross-section, and the mean value was calculated. Another portion of the tissue was fixed in formalin, dehydrated, embedded in paraffin, and sliced. It was then stained with HE and Masson. Non-ablated areas, such as the esophagus, were sampled to assess adjacent tissues. All procedures were approved by the Institutional Animal Care and Use Committee of the General Hospital of Northern Theater Command under the Guide for The Care and Use of Laboratory Animals.

### 2.4 Statistical analysis

Descriptive statistics are expressed as mean ± SD for continuous variables and as frequencies or percentages for categorical variables. A comparison of PFA lesions between groups was performed using an independent-sample t-test. Values of p <0.05 were considered statistically significant. All statistical analyses were performed by using SPSS version 29.0 software (SPSS Inc, Chicago, IL, USA).

## 3. Results

### 3.1 Surgical Procedure

The procedure was completed by PFA in the CVS on 8 healthy canines (with a weight of 35-40 kg and an age of 2.90±0.56 years). The surgery was performed successfully, and the animals survived until the observation time after the operation. The angiography conducted on the CVS provided clear visualization of the GCV, AIV, left ventricular lateral vein, and distal CVS branches (Figure 2A). Guided by a guidewire, the catheter could be delivered into the junction of distal GCV and AIV, as well as the left ventricular lateral vein (Figure 2B, 2C). Following CVS angiography (Figure 2D), the catheter was delivered to AIV (Figure 2E). A clear potential was recorded from the catheter, and the ablation was performed with a biphasic waveform at 1000 V 36 times. The same procedure was employed for the ablation of the distal CMV and GCV. The pulse field generator was observed to be functioning normally throughout the procedure. After the ablation, there was a notable potential reduction, with a 100% acute potential disappearance rate.

**Figure 2.**
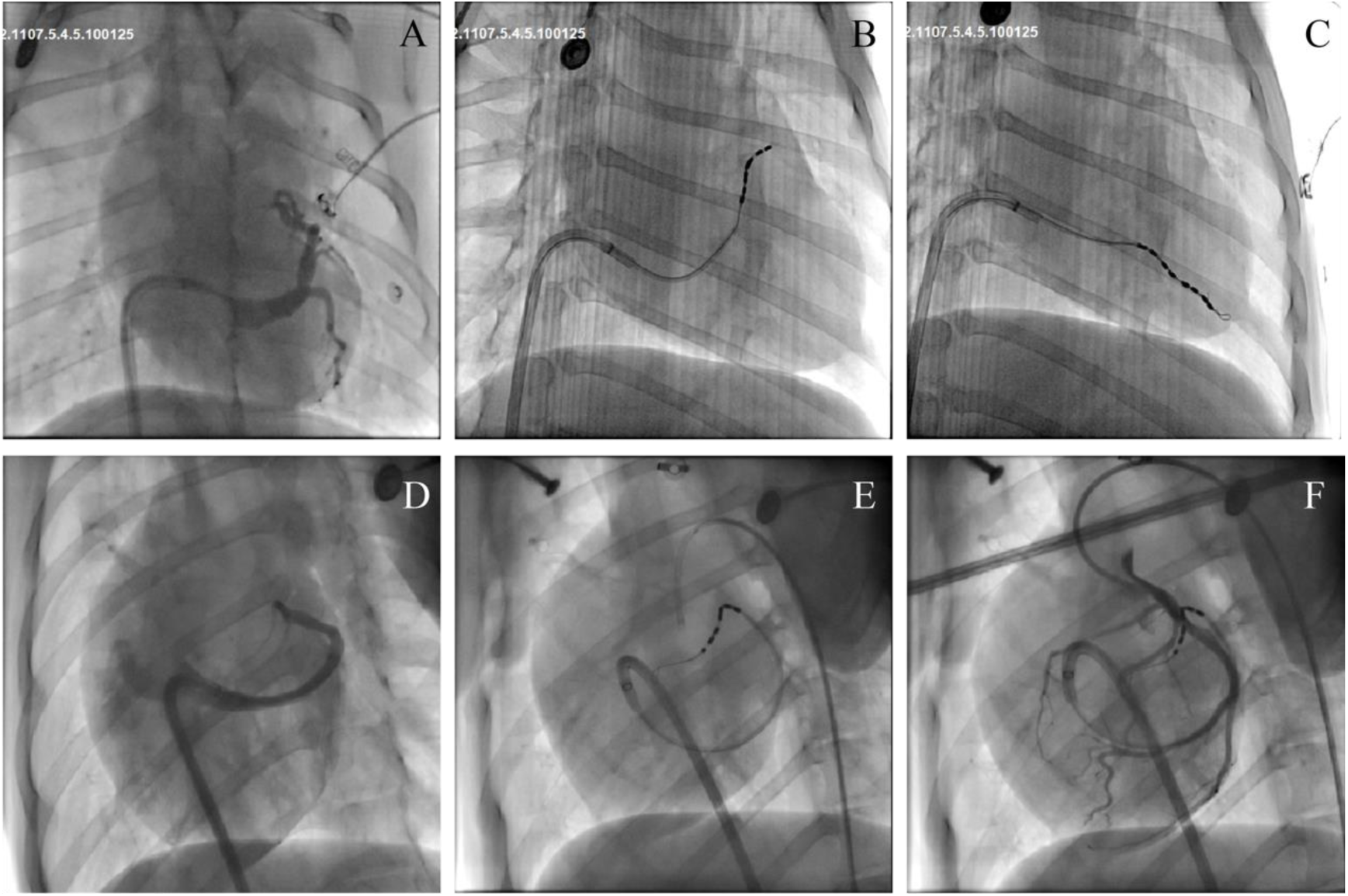
Ablation target shown in the intraoperative DSA images. A: The CVS is displayed (RAO); B: The catheter is located at the junction of the distal GCV and AIV (RAO); C: The guidewire is delivered into distal left ventricular lateral vein; D: The CVS is displayed (LAO); E: The catheter is sent to AIV (LAO); F: postoperative coronary angiography showed the blood flow had no significant changes compared to the preoperative situation (LAO). ABC and DEF are from two different canines.

### 3.2 Gross Observation

Following the euthanasia, a gross and microscopic observation of the heart was conducted. In the 48-hour group, pink bleeding spots were scattered along the vascular zone (Figure 3A), which were not observed in the 30-day group. Grey and elliptical ablation lesions were found to be continuous on each section under TTC staining, perpendicular to the left ventricular lateral vein (Figure 3B). The boundaries of the fat and ablation lesions and the boundaries of the ablation lesions and normal myocardium are discernible (Figure 3C). The size of the lesions resulting from vessels was measured grossly. The mean depth of the lesions in the 48-hour group was 3.92 ± 0.62 mm, while the mean depth of the lesions in the 30-day group was 2.37 ± 0.53 mm. The depth of the lesion in the 30-day group was found to be 39% shallower than that in the 48-hour group (P < 0.001, Table 1). The lesions were shallower when the fat layer was thicker (Figure 3D).

**Figure 3.**
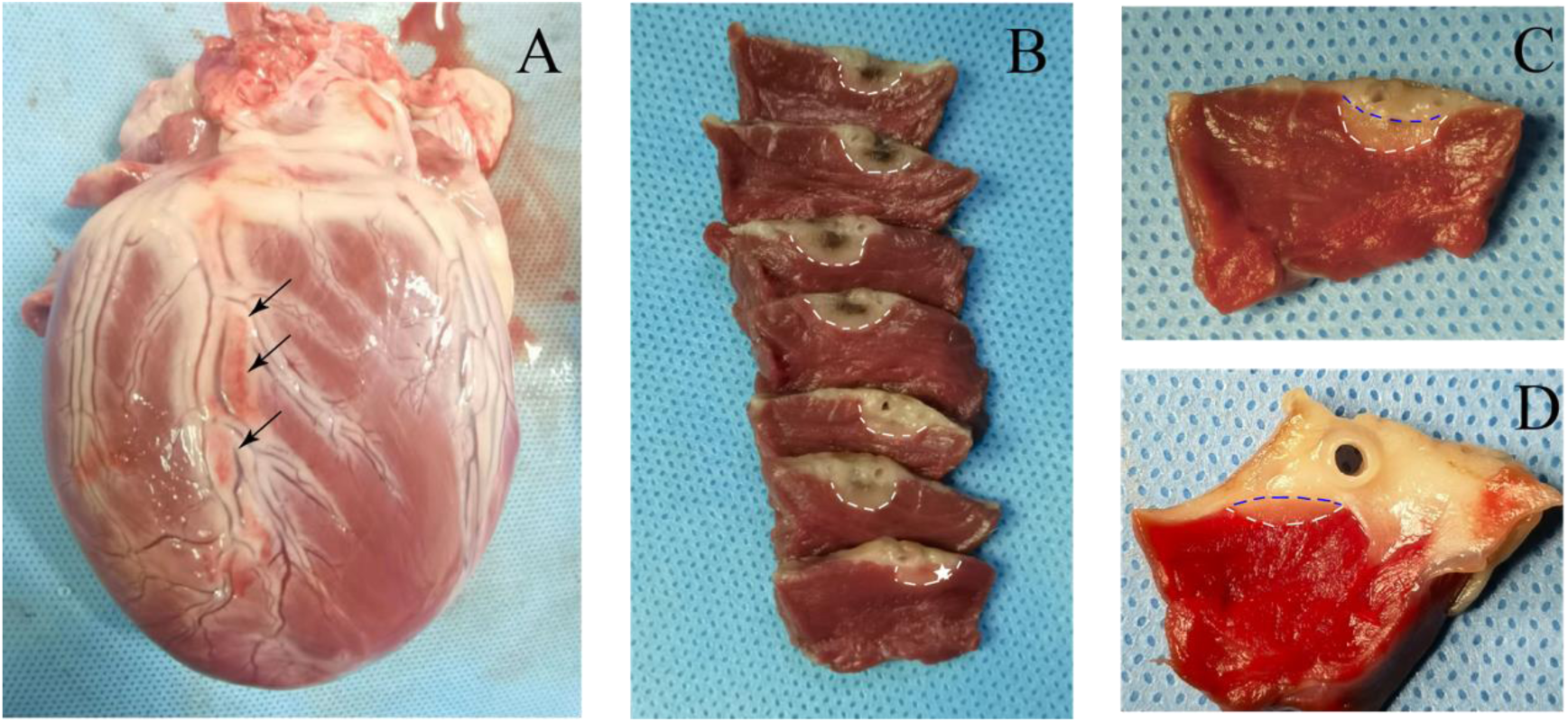
The specimens observed in the 48-hour group. A: Scattered bleeding points (black arrow) can be found along the vascular zone; B: Ablation lesions (white pentagrams) around the left ventricular lateral vein, with clear boundaries with normal myocardial tissue (white dotted line) but unclear boundaries with fatty tissue; C: An enlargement of the tissue marked by the white pentagram in B demonstrates the clear boundaries of the fat-ablation lesion (blue dotted line) and the ablated-normal myocardial boundaries (white dotted line). D: In cases where the fat layer is thicker, the ablation lesion depths are smaller.

**Table 1.**
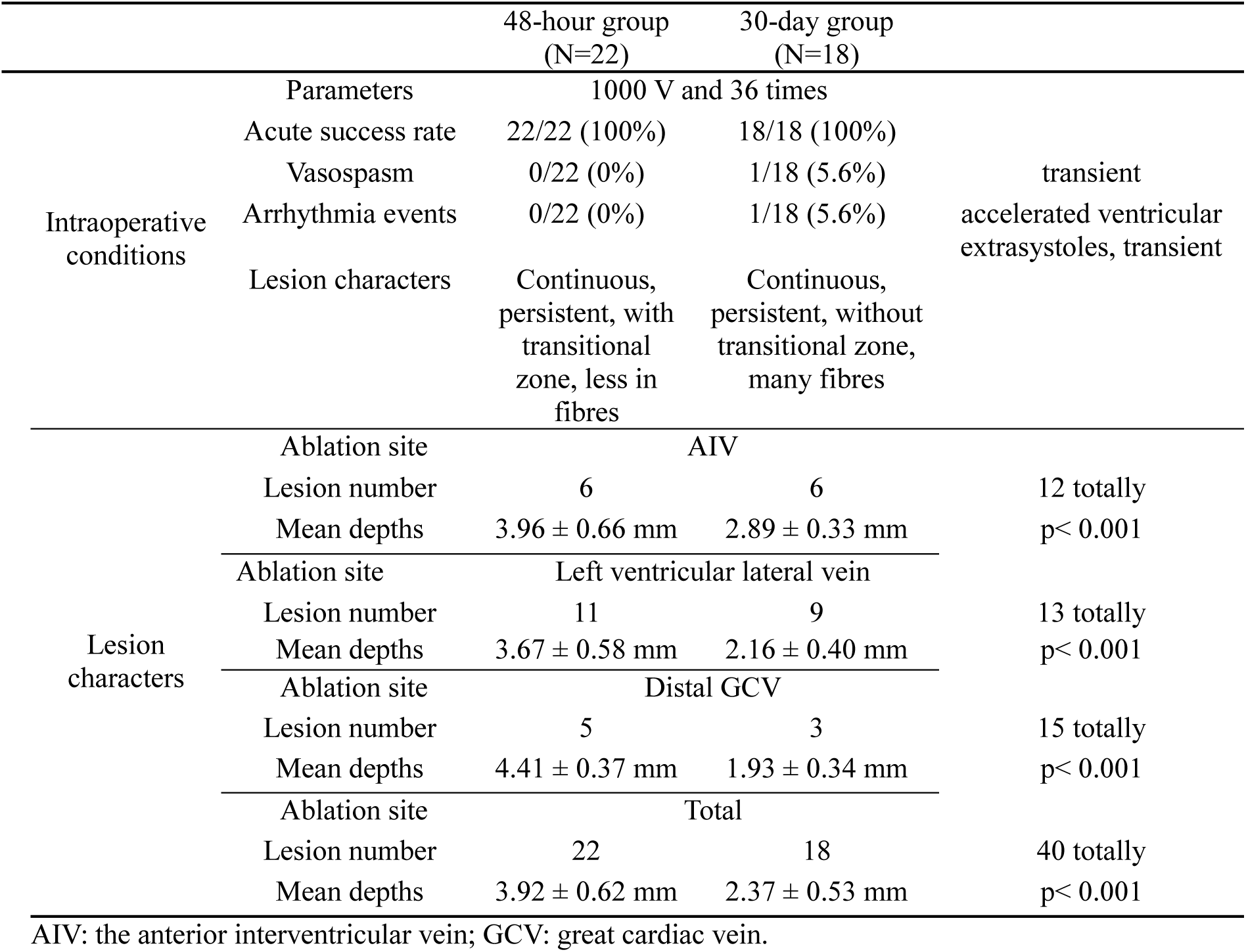
Intraoperative conditions and lesion characters in different groups.

### 3.3 Microscopic Observation

#### 3.3.1 48 Hour-Group

The tissues were obtained from the GCV and CMV (Figures 4A and 4B). Masson’s staining was conducted (Figure 4D, 4E). Both macroscopically and microscopically, pulsed field energy has traversed the fatty tissue, resulting in efficacious ablation lesions on the myocardial surface. The boundary between the injured and uninjured myocardium is delineated (Figure 4F). A gradual transition from the CVS to the normal myocardium can be observed, comprising completely ablated, transitional, and unablated zones. In the center of the completely ablated zone, the myocytes are completely fragmented, the cytoskeleton is destroyed, the arrangement is disordered and sparse, and no obvious fibrosis has formed yet (Figure 4C). In the unablated zone, the cardiomyocyte skeleton was integral, and the cells were arranged neatly and densely (Figure 4F). In the transitional zone, the cells were fragmented but still relatively densely arranged and relatively neat. The boundaries between the different ablation zones were clear (Figure 4F). No obvious constriction, damage, or fibrosis were found at the accompanying coronary arteries.

**Figure 4.**
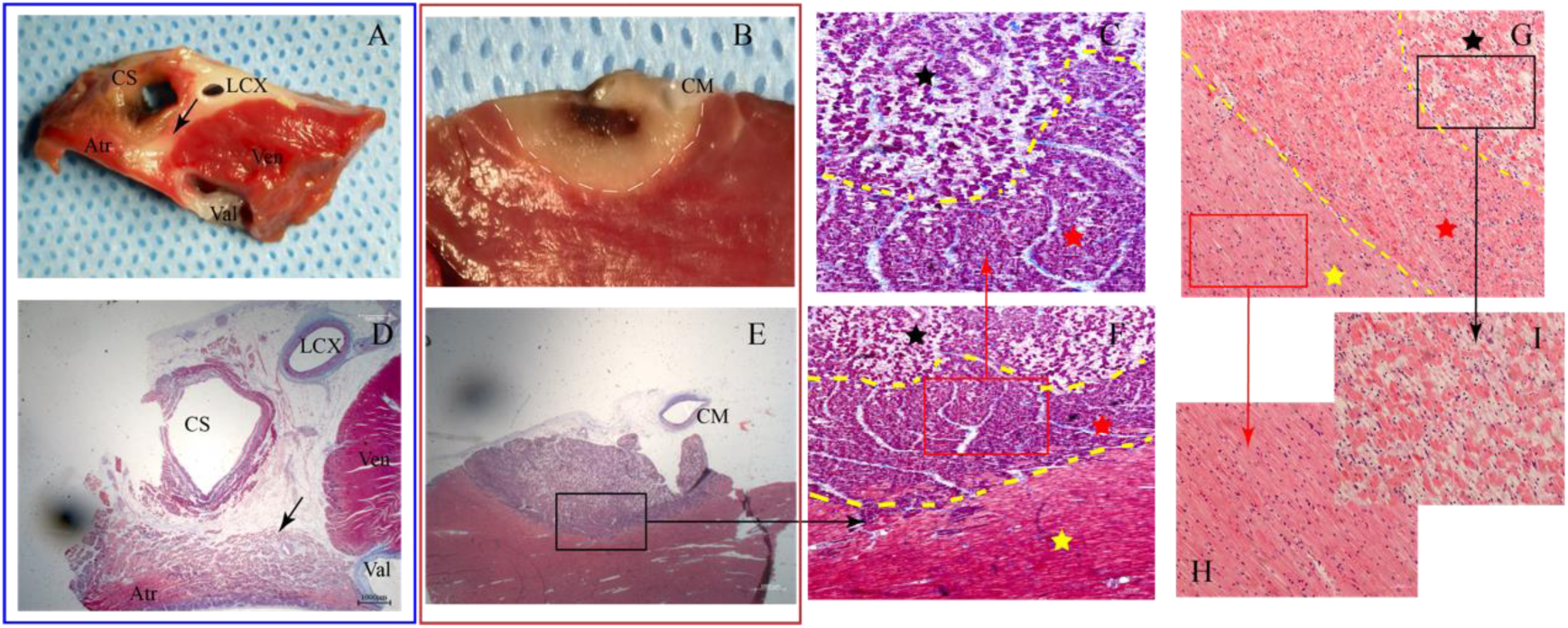
Results of macroscopic and microscopic observations of the 48-hour group. A, D: Gross specimen and corresponding section. The pulsed field energy can pass through the fatty tissue to form an effective ablation lesion. The arrow indicates the ablation lesion; B and E: The ablation lesion from the distal CMV; F is an enlargement of the rectangular area in E, showing the completely ablated zone (black pentagram), the unablated zone (yellow pentagram) and the transitional zone (red pentagram), with clear boundaries (yellow dotted line). The completely ablated zone contains fragmented cells with disordered arrangement, while the unablated zone contains neatly arranged and dense cardiomyocytes. The transitional zone contains fragmented cells but a dense arrangement; C is an enlargement of the red rectangular area in F; G: Completely ablated, transitional and unablated zones under HE staining (black, yellow and red pentagram); H is an enlargement of the unablated zone in the red rectangular area in G; I is an enlargement of the completely ablated zone in the black rectangular area in G; A: coronary artery; V: great cardiac vein; Atr: atrium; Ven: ventricle; Val: valve.

Under HE staining, the complete sequence of ablated, transitional, and unablated zones are also clearly visible (Figure 4G). The completely ablated cardiomyocytes display a range of nuclear shapes and sizes, a swollen cytoplasm, extensive intercellular spaces, and evidence of inflammatory cell infiltration (Figure 4I). The unablated cardiomyocytes are observed to possess integral cytoskeletons, which exhibit the presence of small intercellular spaces (Figure 4H). Cardiomyocytes in the transitional zone are found with swollen cytoplasm and varying nuclear shapes. However, the cells remain relatively closely packed.

#### 3.3.2 30 Day-Group

In the 30-day group, the ablation lesions were still evident (Figure 5A). Under Masson’s staining, the boundary between ablated and unablated areas was clear (Figure 5B, 5C). In the ablated zone, the tissue was homogeneous and slightly transparent, with indistinct cardiomyocyte skeletons. Collagen fibers, which were stained with blue, scattered in intercellular spaces (Figure 5D). In the unablated zone, the cardiomyocytes are observed to be closely arranged, with rare collagen fibers (Figure 5I). Compared with the 48-hour group, no discernible transitional zone was identified. Inflammatory cells can be found in the ablated zone, with deeply stained nuclei. Under the HE staining, cardiomyocytes in the unablated zone were observed to exhibit uniform staining in a light red with scattered nuclei (Figure 6G, 6H). Also, the lesion boundary can be observed.

**Figure 5.**
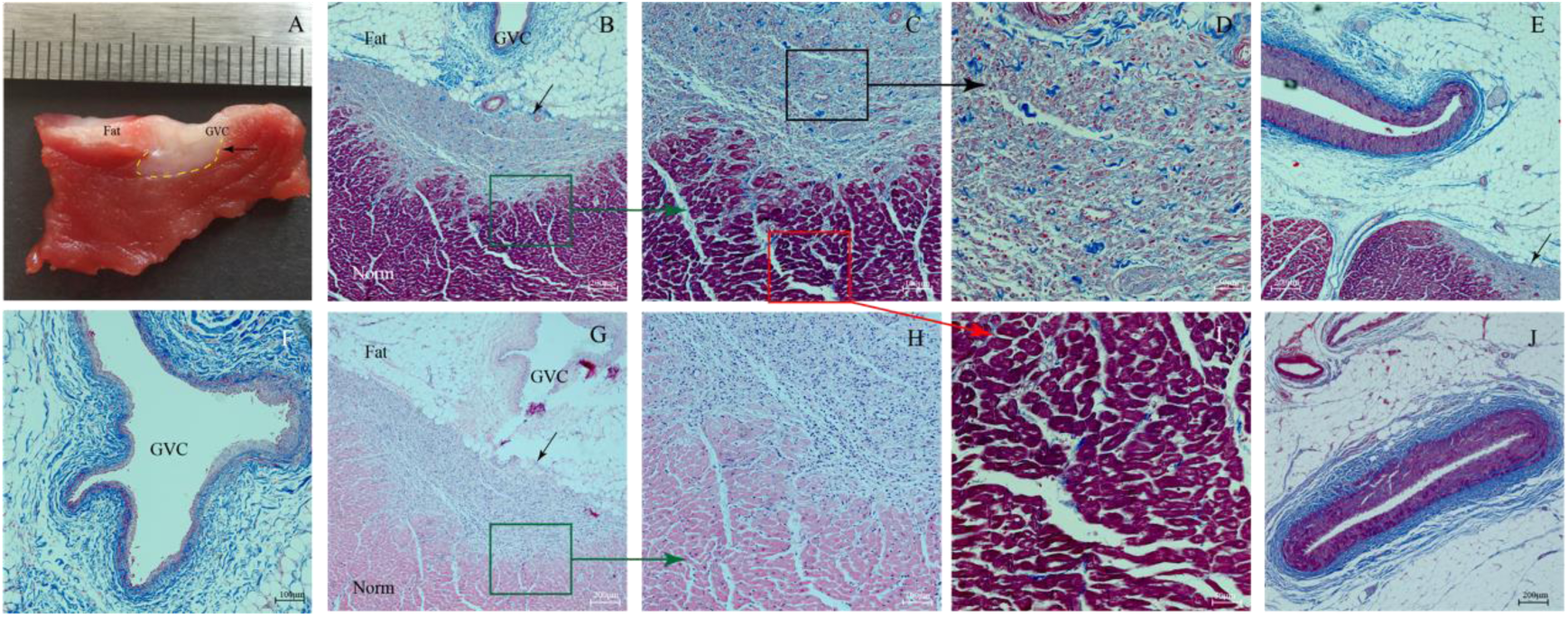
Histological images of the 30-day group. A: Gross observation of the ablation lesion (the black arrow) and the boundary is clear (the yellow dotted line); B: Masson staining of the ablated lesion, the pulsed field energy can penetrate the fat and produce an ablation lesion on the myocardial tissue (black arrow); C: An enlargement of the green rectangular part in B, showing a clear boundary between the ablated and unablated zones; D: An enlargement of the black rectangular area in C, the intercellular spaces are found with collagen fibres; I: an enlargement of the red rectangular area in C, showing the unablated zone; E: no obvious damage or fibrosis were seen in a small artery, which is adjacent to the ablation lesion (black arrow); F: the venous layers where the ablation catheter is located are continuous and intact, no obvious fibrosis, no stenosis or contracture were seen; G: the ablation lesion under HE staining (black arrows); H: an enlargement of the green rectangular area in G, the boundary between the ablated and unablated areas is clear, and no transition zone is found. J: normal small artery; GCV: great cardiac vein; Fat: fat.

### 3.3 Safety

All animals were safe during the operation, and all survived until the observation time point. When pulsed field energy was delivered, the canines had mild skeletal muscle tremors, which were small in amplitude and did not cause displacement during the procedure. Of the 8 canines, one developed coronary artery spasm during pulsed energy delivery and accelerated ventricular extrasystoles were recorded on the electrocardiogram. The operation was immediately stopped, and the guidewire and catheter were withdrawn into the sheath. The angiogram was checked again after 20 minutes, and it found that blood flow had recovered to the pre-ablation state. After ablation, the surface of the catheter poles was carefully checked, and no carbonization was observed. Preoperative and postoperative coronary angiography showed no vasospasm, and the blood supply was unobstructed, with no significant changes compared to the preoperative situation (Figure 2F).

Following the euthanasia, an examination of the adjacent organs was conducted grossly. The surface of the esophagus was observed to be smooth, with no evidence of edema, bleeding, or injury. Within the ablation radius, the adjacent small arteries were observed to be normal by Masson’s staining. Compared with the normal arteries (Figure 5I), the endothelium and the smooth muscle were intact, and no discernible damage was found. No evidence of fibrosis was observed in the entire vascular layer (Figure 5E). The veins where the ablation catheter was located were observed to be continuous and intact (Figure 5F). No evidence of obvious fibrosis, stenosis, or contraction was seen.

## 4. Conclusion and Discussion

This study described a new catheter and methods to treat small vessel-related arrhythmias and verified the efficacy and safety of pulsed field energy application within the distal CVS in a preclinical canine model. The major findings of this study are as follows: 1) The catheter has good maneuverability and precision. Gross examination showed that the ablation lesions formed by the catheter from CVS are effective, continuous, and persistent. 2) In the 48-hour group, the lesion revealed the presence of completely ablated, transitional, and unablated zones. In the 30-day group, the mean ablation depths were found to be 39% shallower than that in the 48-hour group. 3) Histological observation at both 48-hour and 30-day groups showed that the CSV preserved structural integrity and stenosis was not observed in the adjacent artery. Out of a total of 8 canines, a reversible coronary spasm occurred in one during ablation.

Catheter ablation has become the recommended first-line therapy in various clinical guidelines for the treatment of atrial and ventricular arrhythmias^9^. Nevertheless, a part of arrhythmias has a specific target, which can be accessed anatomically closer via small vessel approaches^10^. These targets may be located on the epicardial surface or deeply within the myocardium. Compared with dry pericardiocentesis, the inherent vascular system, comprising the CVS and minor arteries, offers a straightforward route as an additional or alternative approach.

Currently, energies applied in catheter ablation via the CVS involve radiofrequency ablation^11–13^, cryoablation^14^, chemical ablation, guidewire ablation, and pulsed field ablation. However, the catheter cannot reach distal CVS or some small blood vessel branches. Our team has previously reported the use of a guidewire and radiofrequency energy to map and ablate ventricular arrhythmias in the summit region, with subsequent parameter adjustments through animal experimentation^15,16^. However, intravascularly, thermal energy faces challenges because of inadequate power delivery, thermal damage, and thrombus formation. The efficacy of currently available methods for the ablation in small vessels is limited, which restricts their potential for widespread application. In contrast, pulsed field energy, given its tissue selectivity and negligible thermal effects, is a more suitable approach for ablation within the CVS. Buist et al.^17^ first reported pulsed field energy applications within the CVS trunk using a 9 F linear catheter and demonstrated the creation of transmural ablation lesions in atrial myocardium in swine. Then Ivica et al.^18^ and Sevasti-Maria et.al^19^ both reported their application of pulsed field energy within the GCV. Nevertheless, the majority of existing studies are limited to the trunk of CVS because of the limitations of catheter diameter and shape, which prevents them from accessing the smaller branches of the CVS and thus may hinder the treatment of small vessel-related arrhythmias.

The 3 F 10-pole hollow mapping and ablation catheter in the study, with a design of highly maneuverability and flexibility, is designed with a hollow cavity that allows the guidewire to be used for the super selection of vessels and as the guidance to deliver the catheter to the small distal branches of vessels. The catheter can be used for multi-polar synchronous mapping in the CVS, and once the ideal potential is mapped, the direction of the current can be reversed immediately to generate the pulsed field energy between adjacent electrodes near the ideal target, increasing the ablation precision. Unlike traditional thermal-related energy sources, pulsed field energy can pass through fatty tissue to cause effective, continuous, and lasting ablation lesions. We have improved the setting of the catheter parameters and verified their efficacy and safety by animal experiments.

The catheter can cause ablation lesions just as in other PFA studies. The 48-hour group exhibited a transitional zone that had not yet been completely injured under microscopic examination^20^. Also, the ablation lesions in the 30-day group are approximately 39% less in deep compared with those in the 48-hour group, which may be attributed to a combination of factors, including the reversible electroporation formation at the attenuated electric field edge, the regression of inflammatory edema, and the dehydration of scar tissue. Additionally, in our study, the ablation lesions beneath the fat layers were found to be relatively shallow compared with those without fat tissues. These findings align with those reported by Alessio Gasperetti et al.^21^, which may be because of the relatively poor electrical conductivity and potentially influence the distribution of the pulsed field and thus diminish the PFA effect on the surrounding myocardium.

When pulsed field energy is delivered to the ventricular myocardium, the most easily induced and fatal complication is ventricular fibrillation. Thus, pulsed field energy should be administered synchronized with R-wave in this study. Additionally, it is acknowledged that ablating near the coronary artery faces challenges because of the high risks of artery stenosis or occlusion. Animal studies showed a lack of significant injury to vascular by PFA but reversible coronary spasm. In our study, despite a mild edema and small micro-bleedings into the fatty tissue surrounding the PFA application, no visible thermal damage, occlusion, or thrombus of the GCV was observed. No obvious damage was found in the surrounding tissues, such as the esophagus.

Intravascular PFA has been demonstrated to possess certain clinical versatility. In addition to its application in the treatment of arrhythmias, it has also emerged as a promising approach in the interventional management of hypertrophic obstructive cardiomyopathy. At present, radiofrequency and chemical ablation of the interventricular septum are the primary treatments employed^22–24^. However, uncontrollable leakage or incorrect injection of alcohol may result in severe peripheral myocardial necrosis, while the traditional radiofrequency catheter is unable to enter the distal interventricular branch or to be successfully discharged. The 3 F pulse field energy catheter has been initially validated in this study as an effective ablation device in the CVS system. In comparison to alcohol ablation and radiofrequency ablation, it is a safer option. However, its tissue selectivity may result in an inability to damage the supplying artery effectively. Further experimentation is required to confirm whether the device and methods are capable of directly ablating ventricular muscle without impairing small arteries. Concerning safety, the occurrence of reversible arterial spasms, the conduction system injury, and the induction of intraoperative ventricular arrhythmias require further confirmation.

It should be noted that the present study has certain limitations. Firstly, the efficacy and safety of the catheter were only validated in canines in the study, and there is a certain discrepancy between animal and human physiology. Secondly, the study has only verified myocardial ablation in CVS. It is unclear whether the findings can be directly extrapolated to the coronary artery system and the impact of the catheter on coronary artery blood flow. Thirdly, because of the difficulty of constructing an animal model of small vessel-related arrhythmias, we used healthy canines in this study. Using grossly measured ablation depths and histological observation to evaluate efficacy may have certain limitations.

## Data Availability

Data are available within the article or its supplementary materials. The authors confirm that the data supporting the findings of this study are available within the article or its supplementary materials.

## Acknowledgement

The authors would like to acknowledge the technical and production support provided by Shanghai MicroPort EP MedTech Co., Ltd. for the catheter and the pulsed field generator used in this study.

